# Molecular Surveillance of malaria in Nigeria reveals expansion of chloroquine-sensitive infections

**DOI:** 10.1101/2025.07.31.25332537

**Authors:** Olusola Ajibaye, Mary Oboh-Imafidon, Olumide Ajibola, Fiyinfoluwa Olusola, Chinedu Ogbonnia Egwu, Brandon Amambua-Ngwa, Fadel Mouhamadou Diop, Nuredin Mohammed, Umberto D’Alessandro, Catherine Falade, Martin Meremikwu, Adeola Y. Olukosi, Eniyou Cheryll Oriero, Alfred Amambua-Ngwa

## Abstract

**Background:** The success of malaria elimination in sub-Saharan Africa rests largely on the sustained efficacies of the control and prevention chemo-interventions used in malaria treatment and chemoprophylaxis in the highest-burden countries. Data on the impact of these interventions can guide policy for sustained malaria control towards elimination. We report the complexity of infection and genetic diversity of *Plasmodium falciparum* in Nigeria and their antimalarial resistance profiles.

**Methods:** We used targeted amplicon sequencing techniques to analyse 895 *Plasmodium falciparum* clinical isolates collected from the six geopolitical zones of Nigeria, mostly between 2017 and 2021. Genotypes determined from 101 single nucleotide polymorphisms (SNPs) and 36 non-synonymous amino acid mutations associated with antimalarial drug resistance were used to determine the Complexity of infection (COI), drug resistance haplotypes and genetic diversity of the parasites.

**Results:** Overall, there was a high complexity of infection, with 30.1 – 31.3% of the infections having more than a single genetically distinct parasite clone. 55.1% of the parasite isolates carried wildtype alleles of the chloroquine transporter gene, *Pfcrt* codons 74 – 76. The chloroquine-resistant haplotype CVIET was in 26.5% as mono-infections and 14.4% as mixed infections, with an overall presence of 40.9%. High frequencies of sulfadoxine-pyrimethamine (SP) resistance at *Pfdhfr* loci (50%) were observed and the *Pfdhps* A437G mutation was detected in 87.3% of infections. Wild-type haplotypes for *Pfkelch13* were detected, although 12 non-synonymous mutations, were observed.

**Conclusion:** Sustained molecular surveillance for malaria in high-burden countries can support the implementation of current and new malaria strategies for malaria control and elimination.

## Introduction

Since 2020, the global estimates of malaria cases have increased steadily, with approximately 89.7% of the increase occurring in the WHO African Region [1]. In 2023, Nigeria contributed approximately 1.4 million cases to the global increase estimates, being one of the five countries accounting for over half of all cases (25.9%) and deaths (30.9%) [1]. The risk of malaria transmission exists throughout the country, all year round. However, the incidence of malaria is highest in the North, especially the North-eastern parts of the country [2]. This underscores the need to have Nigeria in focus for control strategies to improve the global fight against malaria.

Chemotherapy remains one of the main intervention tools for malaria control in Nigeria, including chemoprophylaxis and therapeutics [3]. A major threat to global malaria control and elimination efforts is the increasing spread of antimalarial drug resistance, including resistance to artemisinin derivatives and partner drugs in first-line Artemisinin-based Combination Therapies (ACTs) [4, 5]. As in most African countries, Chloroquine was used as the first-line treatment of malaria in Nigeria until 2005, following the World Health Organization’s (WHO) recommendation to change to artemisinin-based therapies, due to increasing chloroquine-resistant parasite populations in circulation [6]. Sulphadoxine-pyrimethamine (SP), is the conventional chemoprophylaxis used in intermittent preventive treatment of malaria in pregnancy (IPTp), and with amodiaquine in infants (IPTi) implemented as seasonal malaria chemoprevention in the Northern states [7]. Resistance to these malaria prophylactic drugs has been described in Nigeria, though at low frequencies and Artemisinin combination therapies (ACTs) remain efficacious in Nigeria [8–10]. Currently, malaria chemo-prevention strategies in Nigeria prioritize Intermittent Preventive Treatment in pregnancy (IPTp) and seasonal malaria chemoprevention (SMC) and are in the process of implementing Perennial Malaria Chemoprevention (PMC) and Intermittent Preventive Treatment in school children [7], all of which have SP as the major prophylactic component.

Molecular and phenotypic data on antimalarial resistance in Nigeria is surprisingly sparse despite its high malaria burden. Few studies have reported the prevalence of antimalarial resistance genetic markers and are mostly from the South-west region of the country (Supplementary Table 1). Monitoring trends of antimalarial resistance can be beneficial to assessing the impact of malaria interventions involving therapeutic and prophylaxis agents and to inform policy. With the spread of artemisinin resistance in Southeast Asia [5], emergence and expansion of parasites with delayed artemisinin clearance in East Africa [11–15], there is now more need for improved surveillance, particularly in high malaria-burden regions. Genomic surveillance using advances in deep sequencing technologies not only meets this need but also provides high-resolution information on infection complexity and evolutionary trends that are useful to National Malaria Programs (NMPs) for specific use cases, including the impact of control interventions on transmission.

In this study, we employed targeted deep amplicon to assess and report the frequencies of molecular markers associated with past and current antimalarial drugs across the different regions in Nigeria, showing heterogeneity in the prevalence and connectivity of infections.

## Methods

### Description of samples and ethical considerations

Samples analysed were assessed from different studies carried out in Nigeria (**Table 1**) and sequenced as part of a larger project for the Pan-African Malaria Genomic Epidemiology Network (PAMGEN) and Genomic Surveillance of Malaria in West Africa (GSM). Informed consent was provided by study participants for further genomics analysis of their samples following ethical approvals from the relevant Institutional Review Boards (IRB). IRB approvals were obtained from the Nigerian Institute of Medical Research Institutional Review Board (IRB/21/019; IRB/20/081; IRB/17/038), the Joint University of Ibadan/University College Hospital Ethical Committee (UI/EC/16/0075; UI/EC/19/0114; UI/EC/16/0091; UI/IRC/06/0087), the Oyo State Ministry of Health Ethics Committee (AD 13/479/206), Kebbi State Health Research Ethics Committee (KSHREC 105: 23/2020), the Government of Cross River State Ministry of Health Research Ethics Committee (CRSMOH/RP/REC2017/809; CRS/MH/HREC/017/Vol.V1/109), the Health Research Ethics Committee of the University of Calabar Teaching Hospital (NHREC/07/10/2012; UCTH/HREC/33/489). The PAMGEN and GSM studies received ethical approval from the Gambia Government/MRCG Joint Ethics Committee (Ref: SCC 1626v1.2; SCC 1629).

**Table 1:**
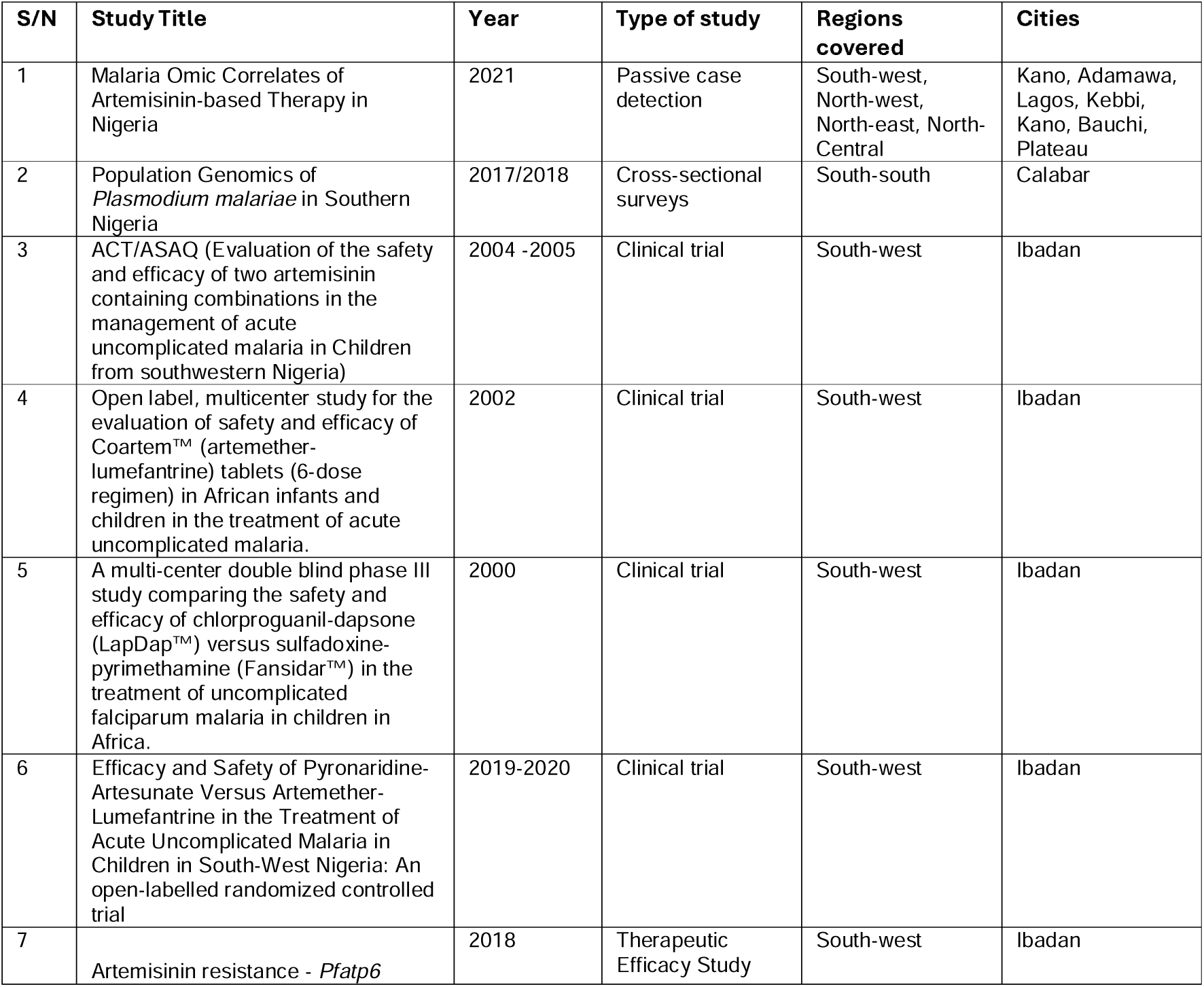
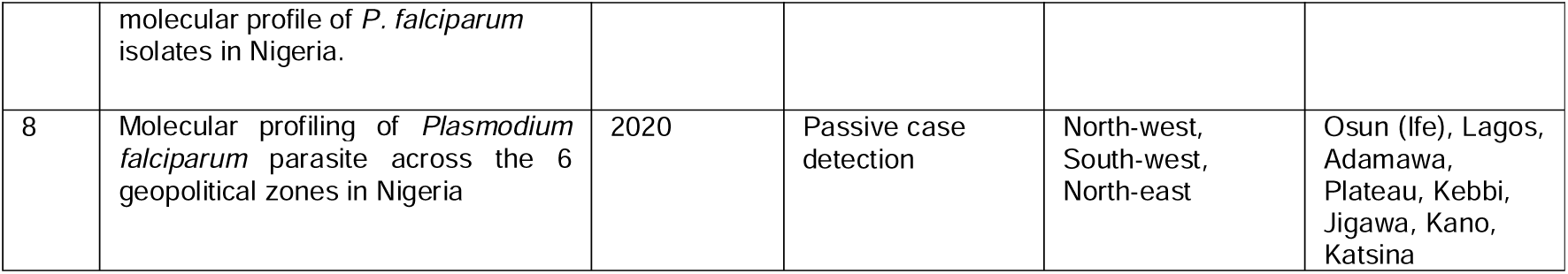
Summary of studies where *P. falciparum* isolates were obtained.

### DNA extraction and confirmatory PCR

Three 6mm spots were punched from the dried blood spots (DBS) for DNA extraction using the automated QIAcube HT according to manufacturers’ protocols (Qiagen GmbH, Hilden, Germany). The confirmation of malaria parasite DNA in the samples was done using an ultra-sensitive qPCR protocol targeting the *var* gene acidic terminal sequence (*var*ATS, 59 copies per genome) on the Biorad CFX96 real-time PCR machine (Hofmann et al., 2015). Samples with Ct threshold less than 40 were taken forward for targeted amplicon sequencing.

### Targeted amplicon sequencing

Selective whole genome amplification (sWGA) was performed to enrich for *P. falciparum* DNA using primers selected on the basis of their differential frequency of binding to the desired (*P. falciparum*) and contaminating (human) genomes (Oyola et al., 2016). Amplicon generation was done using the *P. falciparum* Amplicon Toolkit Protocols [16], consisting of a pool of 136 pre-balanced primers, adjusted to compensate for overperforming and underperforming primer pairs. The amplicons were generated in three panels (GRC1, GRC2 and SPEC) for subsequent barcode labelling using custom sequencing tags. Quality check on the libraries generated was done using Tapestation (Agilent, USA) and library quantitation was done using Kapa library quantification kit (Roche, Switzerland). A final dilution of 8 pMol library was denatured and loaded for sequencing on the MiSeq V2 kit (Illumina, USA).

### Data analysis

Custom bioinformatics pipelines developed at the Medical Research Council Unit The Gambia’s Data Science platform were used to analyse the sequence reads to determine drug resistance alleles at the different loci. A total of 36 non-synonymous amino acid mutations associated with antimalarial drug resistance and derived haplotypes were determined. Genotypes were also determined for 101 single nucleotide polymorphisms (SNPs) carefully selected to capture genetic diversity among *P. falciparum* populations. Complexity of infection (COI) and principal coordinate analysis (PCoA) were performed to compare genetic diversity and transmission intensity across geographic regions, providing insights into the population structure of the parasite and potential clustering patterns related to drug resistance markers. THE REAL McCOIL, a Bayesian approach that uses Markov chain Monte Carlo methods to simultaneously estimate allele frequency and COI was applied to additionally assess polygenomicity in samples and malaria transmission (Chang *et al*., 2017). Network analysis of relatedness between *P. falciparum* parasites was generated using Identity-by-State (IBS) matrices computed from the 101 SNP barcodes and visualized using igraph, tidygraph, and ggraph R packages. All other statistical analyses were performed using R (Version 4.2.2), with visualization performed using the ggplot2 package.

## Results

### Distribution of P. falciparum isolates and Complexity of infection (COI)

Sequence reads were generated for *P. falciparum* isolates from 895 malaria cases, distributed across 19 sites in the country. Approximately 55.5% of the parasite isolates sequenced were obtained from the South-western region compared to 7.5% from the South-south, 11.4% from the North-west, 9.5% from the North-east and 16.1% from the North-central regions. Majority of the samples (78.1%) were collected recently between 2017 and 2021 while some of the samples from Ibadan in the South-east were from retrospective clinical trials spanning across 2 decades (from the year 2000). The spatial distribution of complexity of infection (COI) was overall heterogeneous, ranging from 7.7% in Oyo in the South to 54% in Kano in the North-central regions (**Figure 1**).

**Figure 1:**
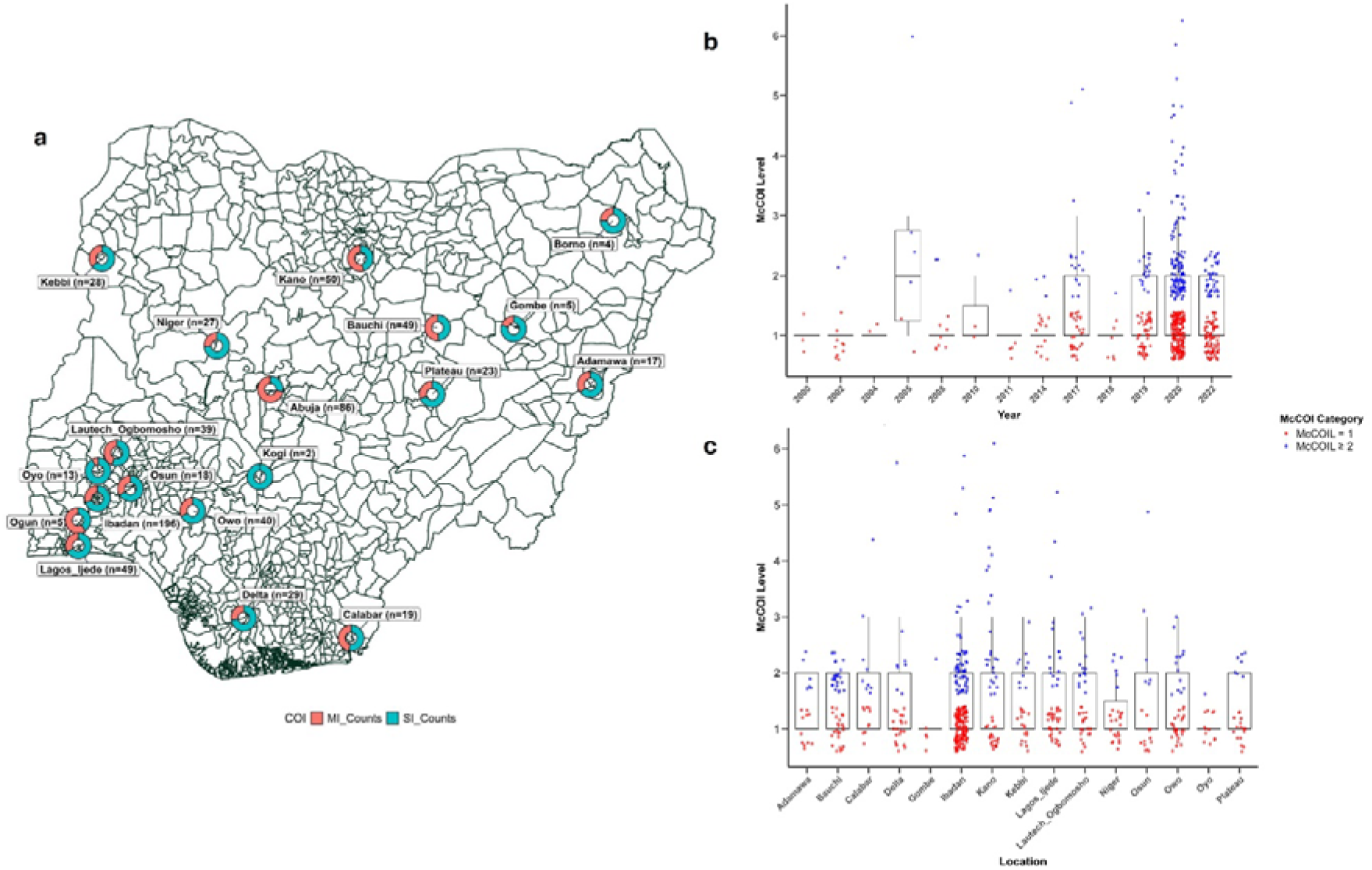
Summary of sample sequenced per site showing a) proportion donut charts of mixed infections (MI) and single infections (SI), and distribution b) by year and c) location (McCOIL = 1 refers to single infections).

Complexity of infection was comparable (30.1 – 31.3% having more than a single genetically distinct clone) between the earlier infections collected in the course of therapeutic efficacy studies from Ibadan and the infections from 2017 to 2021. Infections from Abuja showed an unusual level of heterozygosity and were excluded from subsequent downstream drug resistance genotype analysis, alongside sites with _≤_ 10 parasite isolates sequenced.

### Genetic diversity and relatedness of P. falciparum isolates

A low level of genetic sub-structure was observed among the *P. falciparum* isolates by principal coordinate analysis (PCoA), with two small sub-clusters observed to diverge from the single cluster that included isolates from all sites (**Figure 2a**). Isolates in these subpopulations were from Bauchi and Ibadan. Further exploration by heatmaps of identity-by-state (IBS) pairwise matrix revealed clusters of highly related parasite isolates mostly coming from the same geographic region (**Figure 2b**), suggesting possible clonal expansion of closely related parasites in the population.

**Figure 2:**
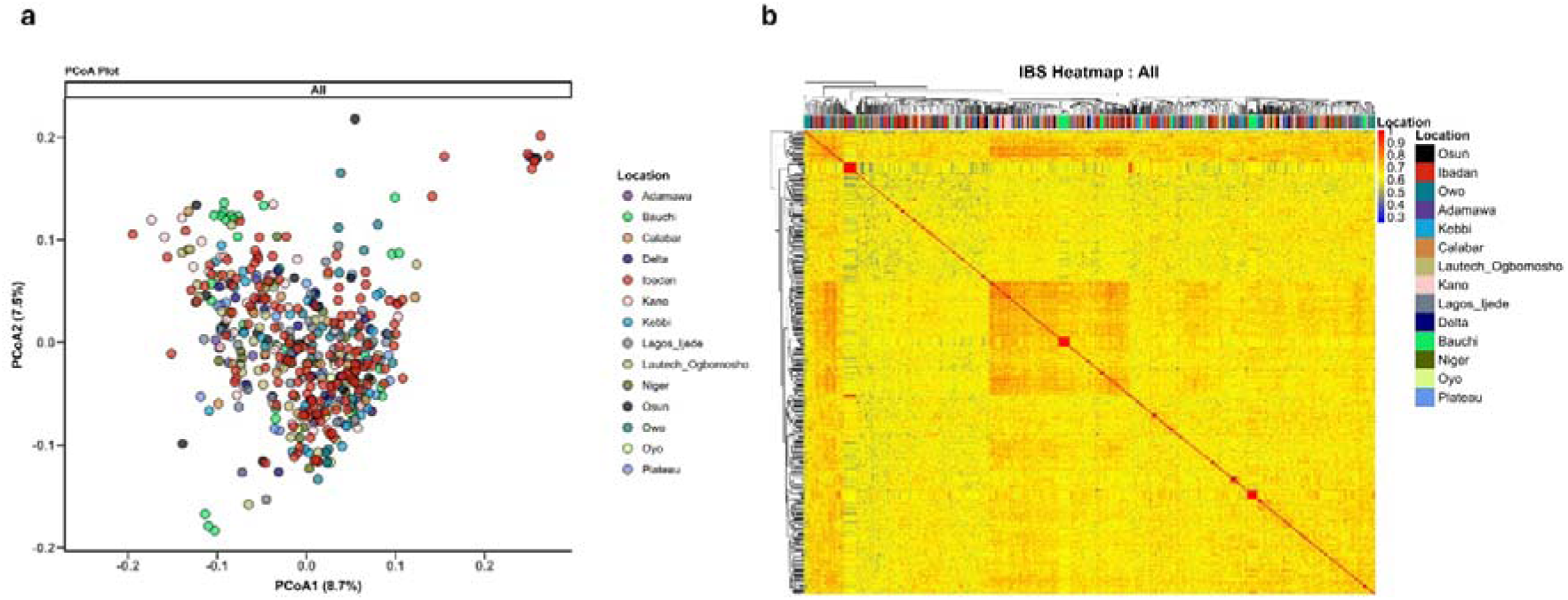
Clustering of parasite isolates showing **a)** genetic diversity using PCoA and **b)** heatmap of identity-by-state (IBS) pairwise matrix.

### Mutational frequencies at Chloroquine resistance markers (Pfcrt)

No mutations were observed at codons 72 and 73 of the parasites analysed for Chloroquine resistance, whereas mutations at codons 74, 75 and 76 were highly similar in frequencies forming the IET haplotype of the *Pfcrt* locus. The frequency of these variants ranged from 5.9% in Kano and Kebbi to 61.9 and 66.7% in Calabar (**Table 2**). Overall, 55.1% of the parasite isolates carried wildtype alleles at *Pfcrt* codons 74 – 76, the proportion was increased to 86.8% in samples collected between 2017 to 2021. The drug-resistant haplotype CVIET was circulating in 26.5% of mono-infections and 14.4% in mixed infections, with an overall presence of 40.9%. (**Figure 3a**). Approx. 4% of samples failed genotyping for this target

**Figure 3:**
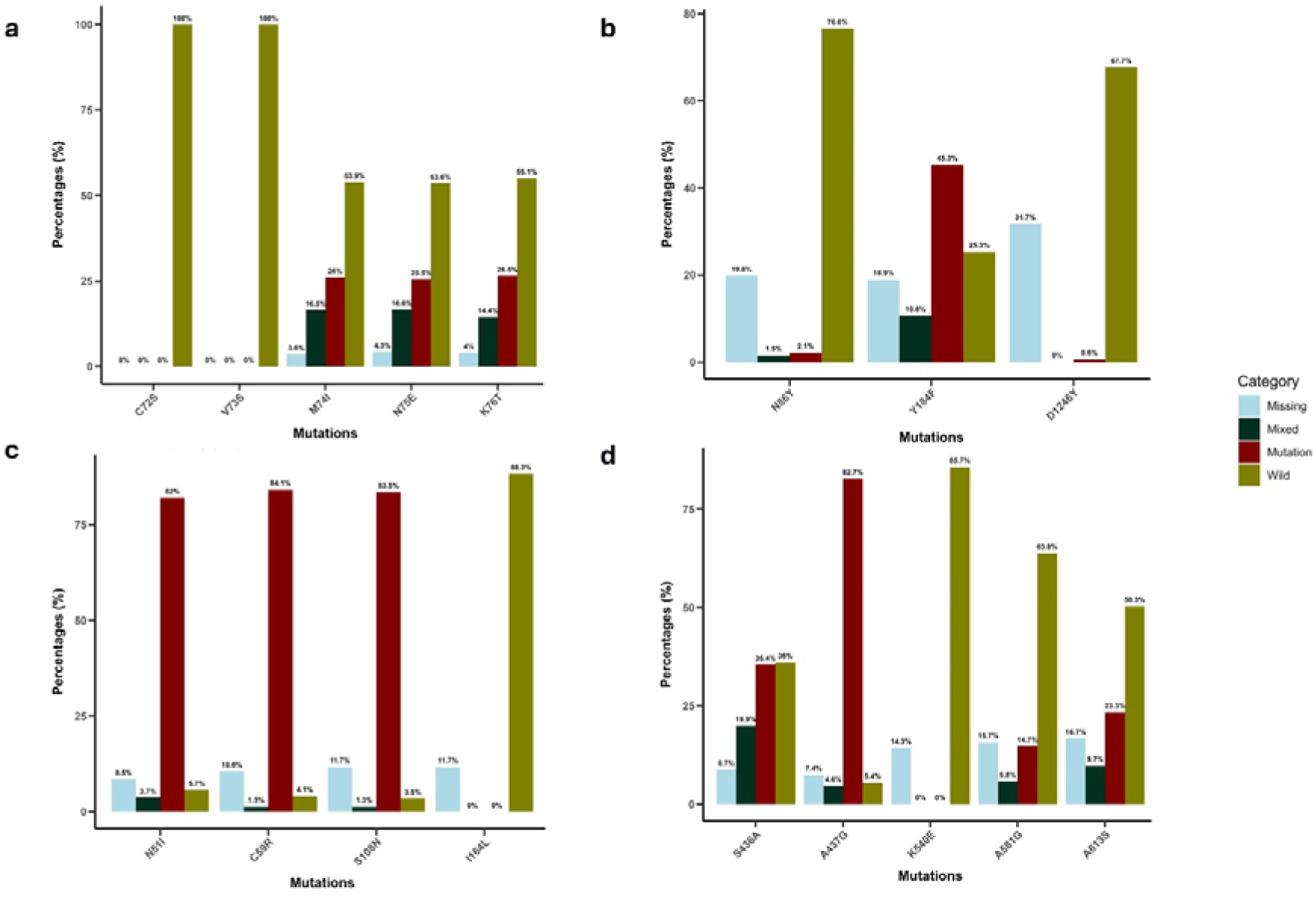
Bar plot of mutational frequencies across the different drug resistance loci **a)** *Pfcrt,* **b)** *Pfmdr1,* **c)** *Pfdhfr*, and **d)** *Pfdhps*.

**Table 2:**
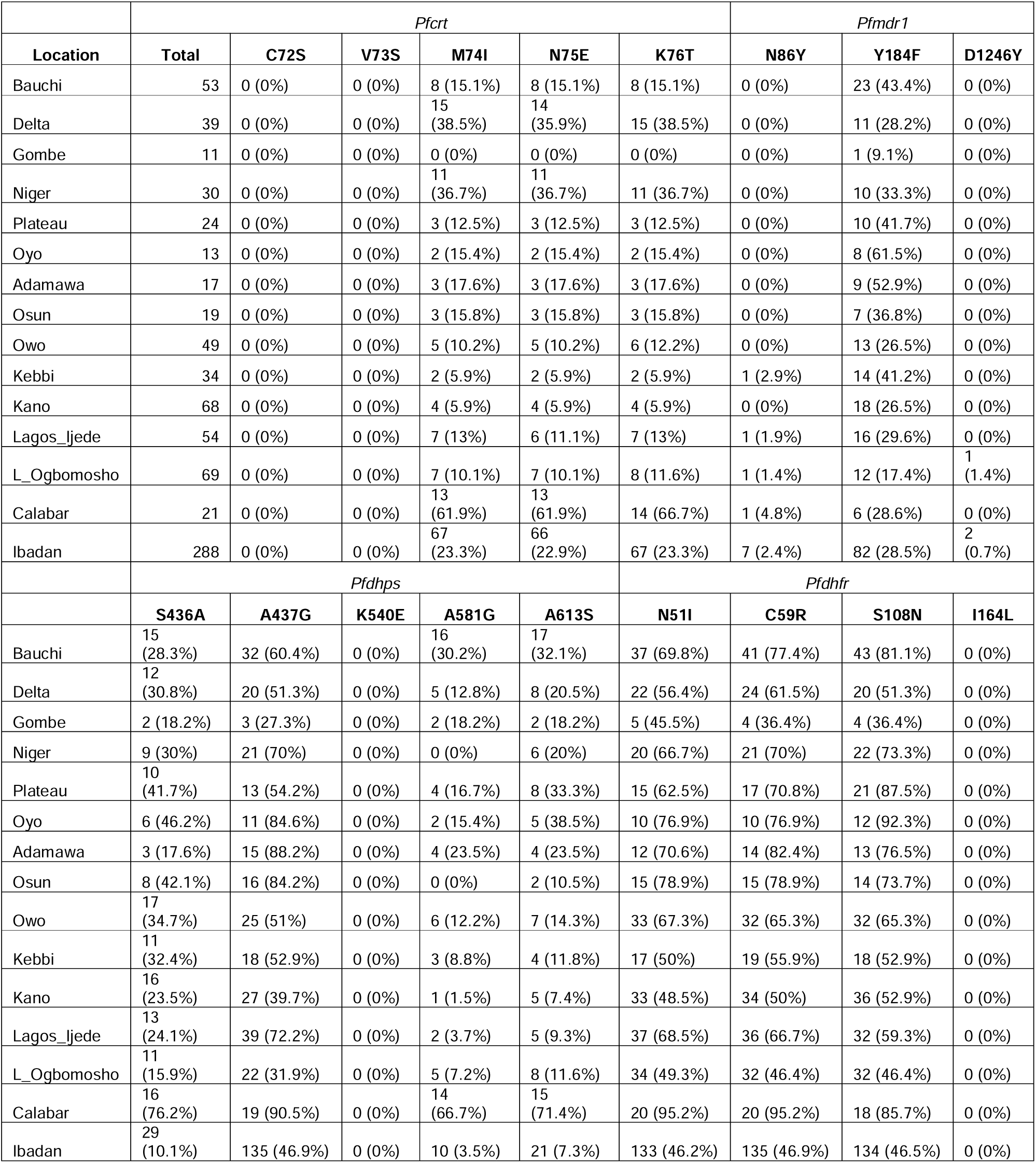
Frequency of drug resistance mutations per site.

### Mutational frequencies at the multidrug resistance gene 1 (Pfmdr1)

Mutations at codons N86Y, Y184F and D1246Y of the *P. falciparum* multidrug resistance gene 1 (*Pfmdr1*) were genotyped. While resistance mutations at codons 86 and 1246 were below 5% across all sites, the frequency of mutants at codon 184 ranged from 9.1% in Gombe to 61.5% in Oyo (**Table 2**). Approx. 18.9% of samples failed genotyping for this target, and a combined resistance frequency of 55.9% was observed for codon 184, compared to 25.3% of wildtype infections (**Figure 3b**).

### Mutational frequencies at sulphadoxine-pyrimethamine resistance markers

Five loci were genotyped at the *Pfdhps* marker for sulphadoxine resistance – S436A, A437G, K540E, A581G, and A613S; and four genetic variants were genotyped at the *Pfdhfr* genetic marker of pyrimethamine resistance – N51I, C59R, S108N, and I164L. High frequencies of *Pfdhfr* mutants (_≥_50%) were observed in parasites from 80% of the sites (**Table 2**), whereas there were no mutations seen at codon 164 (**Figure 3c**). For *Pfdhps*, the A437G mutation was the most prevalent and it was detected in 87.3% of infections, a high proportion of which were mono infections and only 5.4% were wildtype infections (**Figure 3d**). The S436A mutant was seen in 35.4% and 19.9% of mono and mixed resistance infections, respectively. 36% were wildtype at this codon and 8.7% failed genotyping for this target. A relatively lower combined frequency of mixed and mono-resistant infections was observed at codons A581G (20.5%) and A613S (33%) while no mutant at codon K540E was detected.

### Mutational frequencies at artemisinin resistance markers

Wild type (WT) haplotype at the kelch13 BTB/POZ and propeller domain were detected in 95% of monoclonal isolates successfully genotyped. Overall, 12 non-synonymous Kelch propeller domain mutations were observed in 19 samples (**Table 3**). All the variants observed were not amongst the WHO-validated or candidate mutations associated with delayed artemisinin clearance. The mutations were observed in both single and multiple clone infections mixed with wildtype alleles and were mostly from the South-west region of the country.

**Table 3:** Summary of non-synonymous Kelch mutations observed.

| S/N | Mutation | Type | # of samples | Site | Region |
| --- | --- | --- | --- | --- | --- |
| 1 | G382E | Homozygous | 1 | Bauchi | North-east |
| 2 | S459L | Homozygous | 1 | Owo | South-west |
| 3 | V487E | Homozygous | 2 | Owo | South-west |
| 4 | WT V520I | Heterozygous | 1 | Plateau | North-central |
| 5 | WT G533S | Heterozygous | 1 | Ibadan | South-west |
| 6 | V534A | Homozygous | 1 | Ibadan | South-west |
| 7 | WT G544R | Heterozygous | 1 | Ibadan | South-west |
| 8 | WT E556K | Heterozygous | 1 | Ibadan | South-west |
| 9 | A569T | Homozygous | 1 | Abuja | North-central |
| 10 | A578S | Homozygous | 5 | Ibadan, Calabar, Kano | South-west, South-south, North-west |
| 11 | Q613H | Homozygous | 3 | Ibadan, Calabar, Plateau | South-west, South-south, North-central |
| 12 | WT G665S | Heterozygous | 1 | Ogbomosho | South-west |

### Genetic relatedness of drug resistance Infections

Network analysis of genetic relatedness revealed evidence of clonal transmission within chloroquine-sensitive parasites compared to chloroquine-resistant parasites. At a high threshold of pairwise Identity-by-state (IBS) (pairwise IBS _≥_ 0.75 and 0.8), clusters of related pairs among chloroquine-sensitive (wildtype) parasites were observed from multiple geographic locations (Bauchi, Owo, Ibadan), while clusters of related pairs observed in SP quadruple mutant parasites were predominantly from the Bauchi (**Figure 4**).

**Figure 4:**
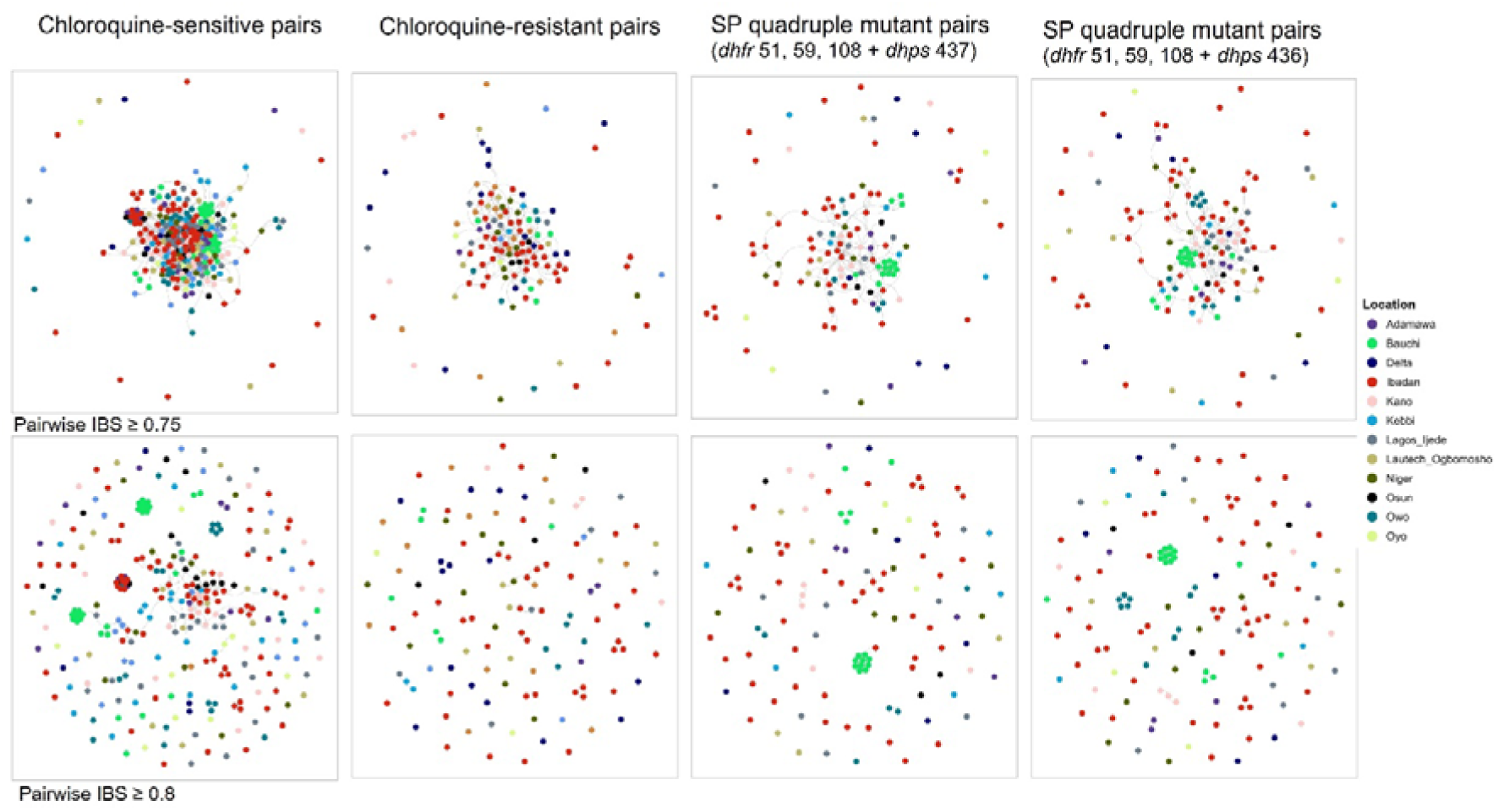
Relatedness network of highly related parasite pairs for chloroquine-sensitive (n = 287), chloroquine-resistant (n = 127) and SP quadruple mutant (n = 122 and 135) parasites sharing pairwise IBS _≥_ 0.75 and 0.8, colours correspond to sampling location.

## Discussion

We report the overall landscape of resistance to past and current antimalarial drugs in Nigeria using high throughput targeted amplicon sequencing. Samples analysed were collected from the different regions in the country, mostly between 2017 and 2021, with one site having temporal samples spanning 19 years (2000 to 2019). Nigeria contributes approximately 25% of the global malaria burden, with chemoprevention and chemotherapy mounting heavy genetic pressure on the parasites. Although there is no confirmed resistance for current first-line ACTs, the use of SP and amodiaquine in chemoprevention warrants the need for molecular and genomic surveillance of legacy resistance to these drugs and to monitor local transmission patterns as intervention strategies are scaled. Overall, we show that complex infections with more than one genetic background remain common across all sites in the country. This data is consistent with the high transmission and aligns with recent reports from a systematic review comparing *P. falciparum* genetic diversity and multiplicity of infection based on *msp-1*, *msp-2*, *glurp* and microsatellite genetic markers in sub-Saharan Africa [17]. Co-transmission of multiple parasites is ubiquitous, and superinfection correlates with entomological inoculation rate, a measure of transmission intensity. Thus, estimates of complex or polyclonal-infection prevalence and population-average COI correlates is an important matrix for tracking transmission intensity and potential indicators of the effectiveness of disease control efforts [18].

Drug-resistant haplotypes for all the antimalarial resistance markers for antifolates, quinolines and artemisinin derivatives were in low frequencies, with exception of the *Pfdhfr* triple mutants which persist above 50%. This pyrimethamine resistance haplotype at *Pfdhfr* is shown to be fixed across most of West Africa [19]. The low frequency of legacy drug-resistant parasites could be a result of the close to two decades of use of artemisinin combination drugs for first-line treatment. The high transmission, genetic diversity and recombination rates in West Africa will result in faster breakdown and replacement of circulating mutant haplotypes in the absence of drug pressure. Moreover, there was heterogeneity in the distribution of markers across regions of the country, which could have been impacted by variance in inconsistent drug pressure, preference for antibiotics for fever treatment by some individuals, use of alternative herbal remedies, and poor quality of antimalarial drugs available through the private sector [20–22].

The low level of *Pfcrt* Chloroquine resistance haplotype was surprising as this differed from those reported in the MalariaGEN Pf7 data release, although the Pf7 had only a small number of infections from Nigeria genotyped from the late 2000s [19]. This reversion to wild variants at *Pfcrt* may have occurred recently. With the withdrawal of chloroquine pressure, *P. falciparum* populations are known to mostly revert to wild-type at *Pfcrt* and some of the variants in *Pfmdr1*. Similar low rates of chloroquine-resistant *Pfcrt* haplotypes have been shown for neighbouring countries such as Ghana, Mali and Burkina Faso, suggesting a trend in West Africa emerging much later than similar reversion in East Africa where chloroquine-sensitive genotypes dominate [23, 24]. However, some of the chloroquine-sensitive infections are clonally expanding especially in urban and semi-urban low transmission regions of the South-west of Nigeria. These present the risk of epidemics and increase pressure from ACT drugs that are readily available in urban centres. As this combination equally presents a risk of other resistance emergence, enhanced molecular surveillance will benefit the National Malaria Programme in designing future intervention strategies, such as multiple first-line therapies to curb the threat of local emergence or establishment of imported resistance to artemisinin combination therapy.

While there were no validated kelch 13 mutations associated with delayed parasite clearance observed in this study, there were cases of non-synonymous mutations observed in monoclonal infections. These variants occurred in domains where validated markers of artemisinin-delayed parasite clearance are known. For example, the E556K mutation observed in this study is close to the WHO-validated P553L mutation, and it is a substitution of an acidic amino acid (Glutamic acid) with a basic amino acid (Lysine), which could have implications for the structure and function of the translated product. Several non-synonymous variants at low frequency have been described across West Africa, but none have been validated as a marker of reduced response to artemisinin derivatives, Nonetheless, their occurrence further emphasizes the need for continuous surveillance and candidate marker validation as a proactive approach towards early warning against resistance to ACTs.

Malaria-endemic countries, which are mostly in Africa, are encouraged to include surveillance in their malaria strategic plans as an intervention. By detecting patterns of resistance that could impact chemoprevention or put current ACTs at risk, these approaches could be more rationally utilised. Moreover, we show the expansion of clonal infection in urban areas, which could result in significant public health issues given the large, interconnected cities in Nigeria. The challenges to the effective delivery of health care to all populations in malaria-endemic countries such as health system weaknesses as well as financial and programmatic factors, highlight the need to develop new and more focused strategies and to ensure that current strategies are better implemented [25]. High-resolution results from genomic surveillance are best suited to support such bespoke intervention strategies.

## Data Availability

All data produced in the present study are available upon reasonable request to the authors

## Financial support

This work was supported by the EDCTP senior fellowship plus to Prof Amambua-Ngwa (Ref: TMA2019SFP-2843). Additional support to the Malaria Population Biology Group at MRCG for molecular epidemiology of malaria was received from the National Institutes of Health Research (NIHR) Global Health (Award ID: 17/63/91 and NIHR134717) and the PAMGEN project, an H3Africa consortium funded by Science for Africa Foundation (H3AFull/17/008).

## Conflict of Interest

The authors have none to declare

## Acknowledgment

We acknowledge all the participants who contributed to the success of this study and all past and current members of the Malaria Population Biology Group at MRCG.

## Notes

### Competing Interest Statement

The authors have declared no competing interest.

### Author Declarations

Nigerian Institute of Medical Research Institutional Review Board (IRB/21/019; IRB/20/081; IRB/17/038) The Joint University of Ibadan/University College Hospital Ethical Committee (UI/EC/16/0075; UI/EC/19/0114; UI/EC/16/0091; UI/IRC/06/0087) The Oyo State Ministry of Health Ethics Committee (AD 13/479/206) Kebbi State Health Research Ethics Committee (KSHREC 105: 23/2020) The Government of Cross River State Ministry of Health Research Ethics Committee (CRSMOH/RP/REC2017/809; CRS/MH/HREC/017/Vol.V1/109) The Health Research Ethics Committee of the University of Calabar Teaching Hospital (NHREC/07/10/2012; UCTH/HREC/33/489) The Gambia Government/MRCG Joint Ethics Committee (Ref: SCC 1626v1.2; SCC 1629).

